# Defining the analytical and clinical sensitivity of the ARTIC method for the detection of SARS-CoV-2

**DOI:** 10.1101/2021.10.09.21264695

**Authors:** Nabil-Fareed Alikhan, Joshua Quick, Alexander J. Trotter, Samuel C. Robson, Matthew Bashton, Gemma L. Kay, Matt Loose, Stefan Rooke, Martin McHugh, Alistair C Darby, Samuel M. Nicholls, Nicholas J. Loman, The COVID-19 Genomics UK (COG-UK) consortium, Samir Dervisevic, Andrew J. Page, Justin O’Grady

**Affiliations:** Quadram Institute Bioscience, Norwich Research Park, Norwich, NR4 7UQ, UK; Institute of Microbiology and Infection, University of Birmingham, Birmingham, UK; Centre for Enzyme Innovation, University of Portsmouth, Portsmouth, PO1 2DT, UK; Hub for Biotechnology in the Built Environment, Department of Applied Sciences, Faculty of Health and Life Sciences, Northumbria University, Newcastle upon Tyne, NE1 8ST, UK; School of Life Sciences, University of Nottingham, Nottingham NG7 2UH, UK; Usher Institute, College of Medicine and Veterinary Medicine, University of Edinburgh, Edinburgh EH9 3JT, UK; Viral Sequencing Service, Royal Infirmary of Edinburgh, Edinburgh, UK; School of Medicine, University of St Andrews, St Andrews, UK; Centre for Genomics Research,University of Liverpool, Liverpool, L69 7ZB, UK; Norfolk and Norwich University Hospital, Colney Lane, Norwich, NR4 7UY, UK; University of East Anglia, Norwich Research Park, Norwich, NR4 7TJ, UK; School of Pharmacy and Biomedical Sciences, University of Portsmouth, Portsmouth, PO1 2DT, UK; School of Biological Sciences, University of Portsmouth, Portsmouth, PO1 2DY, UK

**Author notes:** https://www.cogconsortium.uk (The COVID-19 Genomics UK (COG-UK) consortium). Full list of consortium names and affiliations are in the Supplementary Material.

**Keywords:** SARS-CoV-2, COVID19, Sequencing, NGS, Genome, genomic epidemiology, ARTIC

## Abstract

The SARS-CoV-2 ARTIC amplicon protocol is the most widely used genome sequencing method for SARS-CoV-2, accounting for over 43% of publicly-available genome sequences. The protocol utilises 98 primers to amplify ∼400bp fragments of the SARS-CoV-2 genome covering all 30,000 bases. Understanding the analytical performance metrics of this protocol will improve how the data is used and interpreted. Different concentrations of SARS-CoV-2 control material were used to establish the limit of detection (LoD) of the ARTIC protocol. Results demonstrated the LoD was a minimum of 25-50 virus particles per mL. The sensitivity of ARTIC was comparable to the published sensitivities of commercial diagnostics assays and could therefore be used to confirm diagnostic testing results. A set of over 3,600 clinical samples from three UK regions were then evaluated to compare the protocols performance to clinical diagnostic assays (Roche Lightcycler 480 II, AusDiagnostics, Roche Cobas, Hologic Panther, Corman RdRp, Roche Flow, ABI QuantStudio 5, Seegene Nimbus, Qiagen Rotorgene, Abbott M2000, Thermo TaqPath, Xpert). We developed a Python tool, RonaLDO, to perform this validation (available under the GNU GPL3 open-source licence from https://github.com/quadram-institute-bioscience/ronaldo). Positives detected by diagnostic platforms were generally supported by sequencing data; platforms that used RT-qPCR were the best predictors of whether the sample would subsequently sequence successfully. To maximise success of sample sequencing for phylogenetic analysis, samples with Ct <31 should be chosen. For diagnostic tests that do not provide a quantifiable Ct value, adding a quantification step is recommended. The ARTIC SARS-CoV-2 sequencing protocol is highly sensitive, capable of detecting SARS-CoV-2 in samples with Cts in the high 30s. However, to routinely obtain whole genome coverage, samples with Ct <31 are recommended. Comparing different virus detection methods close to their LoD was challenging and significant discordance was observed.

## Introduction

The COVID-19 pandemic has spread rapidly throughout the world. It began with an unknown case of pneumonia in the city of Wuhan, China (1). The causative pathogen has since been named ‘severe acute respiratory syndrome-related coronavirus 2 (SARS-CoV-2)’. As of October 7th 2021, there have been over 236 million reported cases and 4.8 million fatalities (2). COVID-19 can present with a wide range of symptoms or entirely asymptomatically, making contact tracing and containment difficult based on symptoms alone. Thus, global efforts have focused on widespread testing using diagnostic assays that detect the virus in upper respiratory tract samples (e.g. mouth and nose swabs). A range of diagnostics platforms are now available through different commercial vendors, some with fundamentally different methods for detection.

SARS-CoV-2 tests belong to one of three categories based on what is targeted. Firstly, nucleic acid tests detect the presence of viral RNA; these typically amplify and detect >1 region of the SARS-CoV-2 genome via RT-qPCR or an alternative nucleic acid amplification technology e.g. LAMP. Secondly, antigen detection tests that detect viral proteins typically in upper respiratory tract samples using lateral flow immunoassays. Finally, there are antibody tests that detect host immune responses in blood thereby indirectly detecting SARS-CoV-2. In addition to these methods, targeted and metagenomic sequencing can be used to detect SARS-CoV-2 in patient samples.

SARS-CoV-2 genome sequencing has enhanced contact tracing and containment efforts by providing information on pathogen evolution, population structure and transmission between individuals. There are different protocols for sequencing and analysing SARS-CoV-2 genomes, one of which is the ARTIC protocol (7). The ARTIC protocol was initially released in January 2020 and utilises tiling PCR based genome sequencing (2 × 48plex PCR reactions covering the ∼30kb RNA genome) developed using the Primal Scheme tool (3) and an informatic pipeline refined from previous work analysing viral genomes during the Ebola and Zika outbreaks (4). Amplicon sequencing has been used to sequence more than 96% (n=1560345/1625562) of all the publicly-available SARS-CoV-2 genomes uploaded on the European Nucleotide Archive (https://www.covid19dataportal.org) (accessed 07-10-2021), of which the ARTIC protocol or derivatives of the protocol account for virtually all of these data. Defining the performance characteristics of the ARTIC protocol in comparison to commonly used diagnostics tests enables scientists to select samples that will sequence successfully, providing cost savings. The protocol may also be useful for benchmarking SARS-CoV-2 diagnostic tests.

Here, we define the limit-of-detection (LoD) of the ARTIC protocol using different concentrations of SARS-CoV-2 control material and compare its sensitivity with a number of diagnostic platforms currently in use in the UK National Health Service (NHS). The ARTIC results (COVID-19 positive or negative) were also compared to the result (COVID-19 positive or negative) reported by the originating hospital laboratory. Understanding the boundaries and limitations of the ARTIC protocol will help interpretation of all the data that has been generated so far. We determined the LoD of ARTIC by testing different concentrations of SARS-CoV-2 control material, in triplicate. A set of over 3,600 clinical samples from three UK regions were then analysed to determine the limits of the ARTIC protocol in real-world samples and to compare its performance with twelve clinical diagnostic assays. Finally, we present a bioinformatics tool RonaLDO for assessing and comparing ARTIC sequencing runs.

## Methods

### Mock samples

The limit of detection (LoD) of the ARTIC protocol was determined using different concentrations of SARS-CoV-2 control material that had been quantified by digital PCR. A fixed number of inactivated viral particles (equivalent to 0, 1, 5 10, 50 and 100 genome copies) of Qnostics SARS-CoV-2 Q Control 01 (6) were spiked into viral transport media (200ul per sample) to create mock diagnostic samples. Samples were prepared in triplicate for each concentration and RNA was extracted using the Quick DNA/RNA Viral Magbead kit (Zymo)(7).

### Clinical samples

SARS-CoV-2 positive upper respiratory tract swab samples were obtained from hospital diagnostic laboratories collaborating with the COVID-19 Genomics UK consortium (COG-UK) sequencing study. This included samples from collected from hospitals linked to Quadram Institute Bioscience (NORW), University of Portsmouth (PORT), Northumbria University (NORT) and University of St Andrews (EDIN). For NORW, hospital sites include Norfolk & Norwich University Hospital, Queen Elizabeth Hospital King’s Lynn, James Paget University Hospital, Ipswich Hospital, Colchester Hospital and West Suffolk Hospital. For PORT, hospital trusts include Brighton and Sussex University Hospitals NHS Trust, Portsmouth Hospitals University NHS Trust and University Hospital Southampton NHS Foundation Trust. All samples were collected prospectively as part of routine clinical diagnostic testing and excess sample shared with the sequencing sites under approval by Public Health England’s Research Ethics and Governance Group (PHE R&D Ref: NR0195).

A subset of samples were selected for which both details of the diagnostic test results were included and the sequencing run had been proved reliable. A sequencing run was considered as reliable when: the associated negative controls for that sequencing run had less than 100 total SARS-CoV-2 mapped reads; less than 4% genome recovery (equating to 3 amplicons); and the sequencing run itself encountered no issues impacting the quality of the read data.

### Genome sequencing

cDNA and multiplex PCR reactions were prepared following the ARTIC nCoV-2019 sequencing protocol v2 (8) with version 3 of the primer scheme (https://github.com/artic-network/artic-ncov2019/tree/master/primer_schemes/nCoV-2019/V3). Sequencing was performed using Illumina or Nanopore sequencing platforms.

### Demultiplexing and read mapping

Sequenced read data from NORW included data generated through Oxford Nanopore and Illumina, and were demultiplexed accordingly. Illumina reads from the NORW site were demultiplexed using bcl2fastq2, allowing zero mismatches for indexes. Oxford Nanopore reads were demultiplexed using Guppy with ‘strict’ and ‘HAC mode on. ‘Strict’ required that demultiplexing barcodes were recovered from both ends. All reads were further processed using the ‘ncov2019-artic-nf’ pipeline, a Nextflow pipeline for running the ARTIC network’s field bioinformatics tools (9). The ‘ncov2019-artic-nf’ pipeline was run with ‘--ivarMinDepth 100’ and with all other parameters set to default, as in the repository’s configuration.

Sequenced read data from the PORT site, which were all generated through the Oxford Nanopore platform, were demultiplexed using MinKNOW with ‘--require_barcodes_both_ends’ set. Reads were further processed using the ARTIC network’s field bioinformatics pipeline available through bioconda.

Sequenced read data from EDIN and NORT sites, which were all generated through the Oxford Nanopore platform, were demultiplexed on the GridION with ‘strict’ and ‘HAC mode’ on. Reads were further processed using the ARTIC network’s field bioinformatics tools available through bioconda. The full pipeline is available in (10). The Wuhan-Hu-1 reference genome (accession number MN908947.3) was used throughout for all sites.

### Calculating and comparing mapping coverage

Reads from samples sequenced on the Illumina platform were filtered; when the read length was 150 bp, only reads with a number of mapped bases with greater than 148 base pairs (bp) were included in subsequent analysis. This addressed the issue of samples/reads with potential primer dimer artifacts. Reads from samples sequenced using the Oxford Nanopore Technologies (ONT) platform were filtered using the default parameters of the ARTIC network’s field bioinformatics tools. Mapping coverage and platform comparison analytics were performed using RonaLDO (11) (v1.0.0).

### Data release

Raw sequence data from clinical samples were deposited in and are available from the European Nucleotide Archive under BioProject accession number PRJEB37886. All consensus genomes are available from COG-UK (https://www.cogconsortium.uk) and high-quality genomes are also available from GISAID (5) with individual sample accessions listed in Supplementary Table 1. Sequence data from LoD experiments are available under BioProject accession number PRJEB41469.

### RonaLDO

RonaLDO is Python version 3 software created as part of this study to validate the sequence data and is now available under the GNU GPL3 open source licence from: https://github.com/quadram-institute-bioscience/ronaldo. RonaLDO uses data from a single SARS-CoV-2 sequencing run as input and generates metrics from this; metrics from multiple sequencing events are combined, filtered and plots created.

We developed RonaLDO based on the premise that, for a given sample to be SARS-CoV-2 positive, the sequenced reads would contain a number of reliable reads that map to the SARS-CoV-2 genome. In RonaLDO, the definition of reliable reads is different depending on the sequencing platform used. Reads produced through Oxford Nanopore are considered reliable by RonalDO if they map to the SARS-CoV-2 genome, whereas Illumina reads also require that the number of mapped bases exceed a defined threshold (148 bp by default). This is to account for short reads (often less than 70 bp) within Illumina sequencing data, which are likely to be primer dimer artifacts. In the ARTIC pipeline, the default cutoffs for read length is 400 base pairs for Nanopore and 20 base pairs for Illumina (12). In RonaLDO, the specific number of reliable reads required should exceed an absolute number of reads (30 by default) and be higher than the number of full-length mapped reads in the negative controls (typically caused by barcode crosstalk) done as part of the same sequencing run.

In a preprocessing step, reads from all samples, including negative controls, were aligned to the Wuhan Hu-1 reference genome (accession number MN908947.3), using the ARTIC bioinformatics pipeline, outputting read alignments in BAM format (12). Each sequencing run was specified as input to RonaLDO in the format of a directory of all resulting BAM files, alongside diagnostic platform metadata such as the Ct value. Each analysis run of RonaLDO initially checked the negative controls for significant proportions of SARS-CoV-2 material; if this was the case then the analysis was halted as the run was considered contaminated (laboratory, reagent or cross contamination). If analysis proceeded, then the ‘Calculate’ module calculated quality metrics for the given samples, such as genome coverage, genome recovery, and the number of reads. Once all the metrics were compiled, for both samples and negative controls, they were assessed using the ‘filter’ module which determines whether samples were positive using a number of thresholds, and produced a final results table in text-delimited (csv) format. These thresholds can be changed by the user through a set of command-line parameters at run time. An additional ‘plot’ module was used to produce the charts presented here.

## Results

### Limit of detection (LoD) for the ARTIC protocol

A set of mock samples with fixed concentrations of viral copies (0, 1, 5, 10, 50, & 100) were run through the ARTIC protocol (three biological replicates) and sequenced on an Illumina NextSeq 500. A proportional increase in genome recovery was observed that enabled the percentage of ARTIC amplicon sites with >=10X SARS-CoV-2 coverage to be defined (Figure 1A).

**Figure 1:**
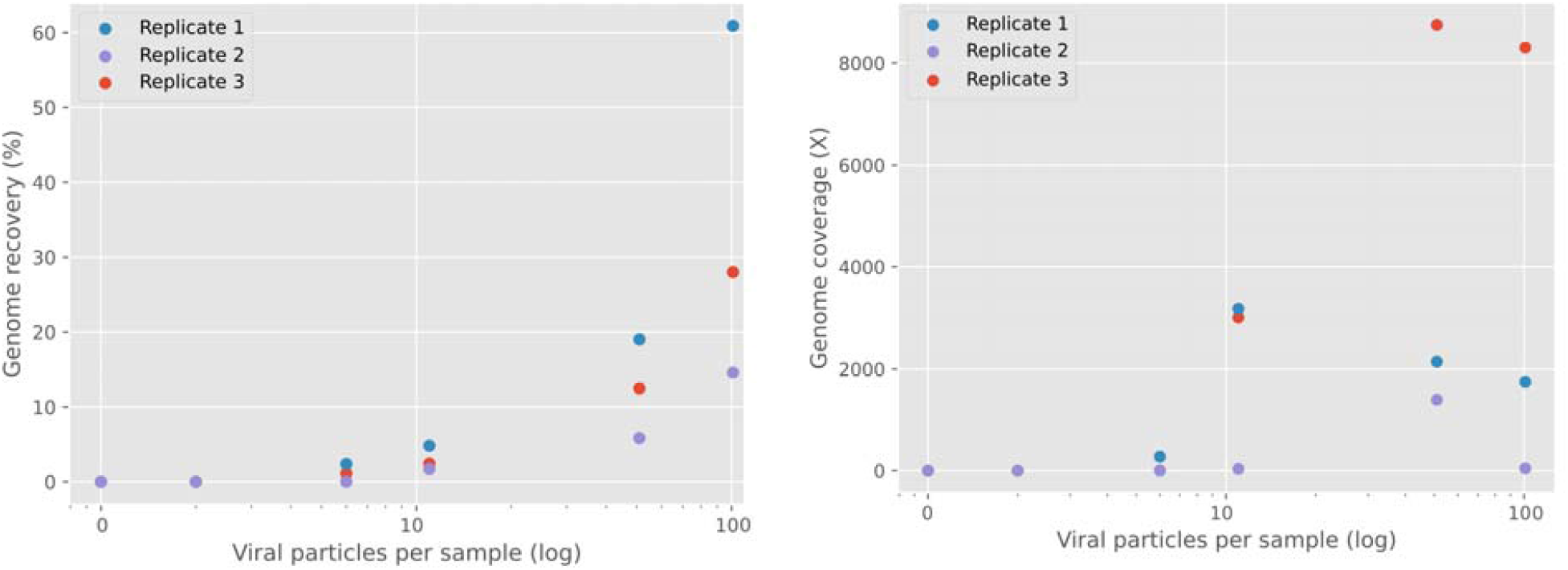
Genome recovery and coverage with control samples. (A) Effect of the number of viral particles per sample on genome recovery (%) (B) Effect of the number of viral particles per sample on genome coverage (Depth of coverage). Replicates are colour coded according to key. Genome recovery was defined as the percentage of SARS-CoV-2 genome sites with greater than 10X coverage.

In two replicates, ARTIC required at least five virions (in 200ul) to recover full mappable sequencing reads, which was greater than 1.14%-2.38% genome recovery. This equates to 1-2 ARTIC amplicons. The third replicate required at least ten virions for 1.7% genome recovery (Figure 1A). Genome depth of read coverage showed no clear relationship with viral copy number (Figure 1B). We therefore define the LoD for the ARTIC protocol to be a minimum of between five and ten virus particles per sample, which equates to 25-50 virus particles per mL. Thus, ARTIC has a comparable sensitivity to the five other diagnostic platforms currently used (Table 1). All currently available diagnostic assays target two regions of the SARS-CoV-2 virus, although the exact targets are not routinely disclosed by the manufacturers. Furthermore, manufacturers measure the LoD in different ways making direct comparisons difficult. Similarly, the LoD can be different for different targets on a single platform.

**Table 1:** Limit of detection (LoD) comparison of SARS-CoV-2 diagnostic systems.

| System (Assay) | Target | LoD/ml of primary sample | LoD/10ul input material | Reference |
| --- | --- | --- | --- | --- |
| ARTIC (V2) | Whole Pathogen | 25-50 copies | 0.9-1.8 copies | This study |
| Hologic Panther (Aptima® SARS-CoV-2 Assay) | ORF1ab (2 targets) | 62.5-125 copies | 7.4-14.8 copies | (13) |
| AusDiagnostics (SARS-CoV-2, Influenza & RSV 8-WELL) | ORF1 | 2150-4325 copies | 86-173 copies | (14) |
|  | ORF8 | 175 copies | 7 copies |  |
| Cepheid Xpert Xpress (SARS-CoV-2/Flu/RSV) | N2 | 0.005 pfu | 0.00005 pfu | (15) |
|  | E | 0.020 pfu | 0.00020 pfu |  |
| Roche Cobas (cobas® SARS-CoV-2) | ORF1ab | 0.009 TCID50/ml | 0.00009 TCID50/ml | (17) |
|  | E | 0.003 TCID50/ml | 0.00003 TCID50/ml |  |

ARTIC also demonstrated high sensitivity when used on clinical samples. By comparing reported cycle threshold (Ct) values from clinical samples with the resulting genome recovery when sequenced, there was a decrease in genome recovery in samples with a Ct higher than 30 (Figure 2), where a base in the genome is said to be recovered if it has reads covering at least 10X for Illumina data and at least 20X for Nanopore data (the minimum depth required to call a variant). Despite this, higher Ct samples (33-38+) had at least 4% genome recovery, which was still higher than the associated negative controls. So, while high Ct samples were not appropriate for phylogenetic analysis, there was evidence that ARTIC could detect presence or absence of SARS-CoV-2 in high Ct samples.

**Figure 2:**
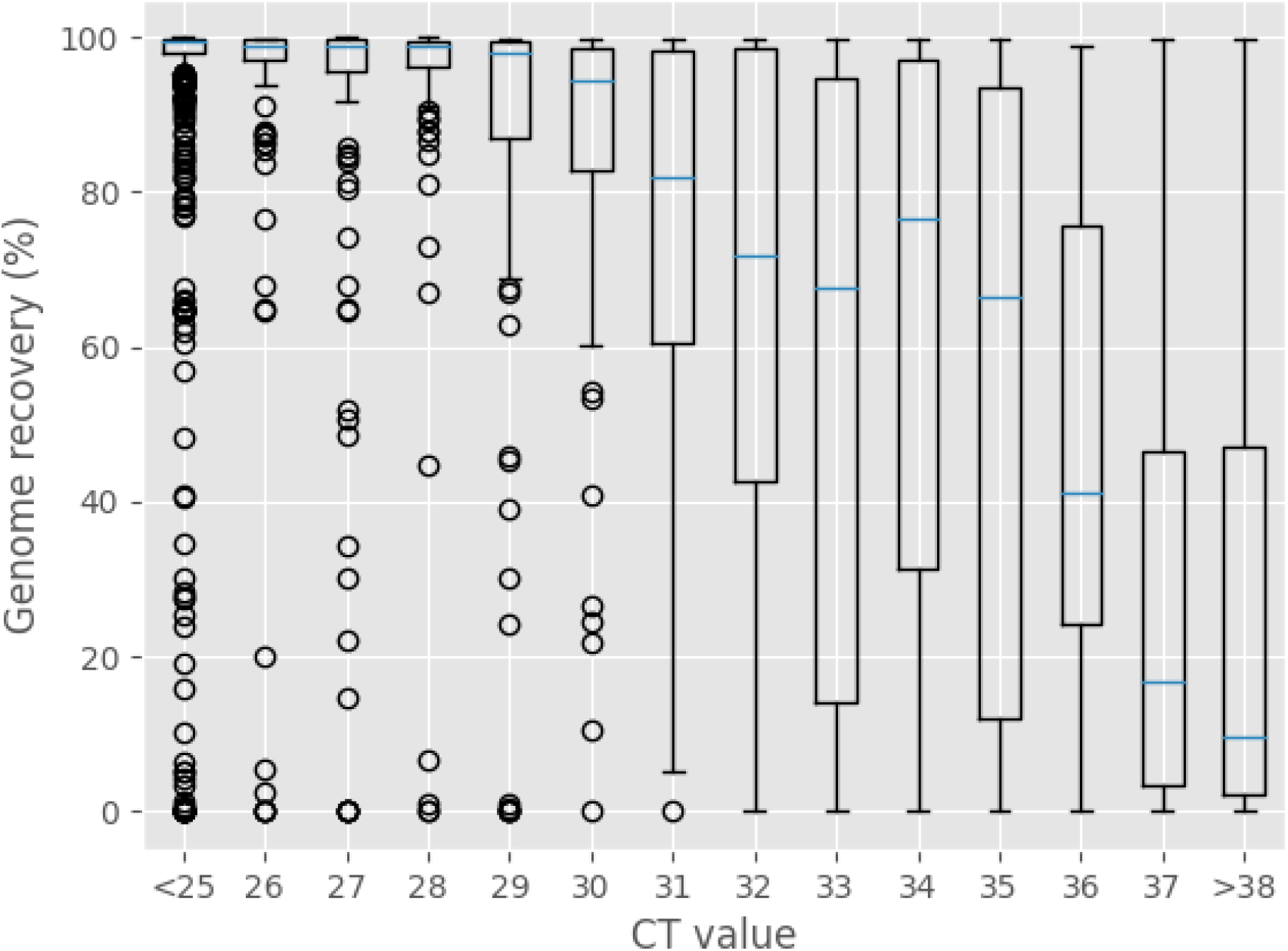
Cycle threshold (Ct) vs genome recovery from sequenced COG UK data. Comparison of the cycle thresholds (Ct) and resulting genome recovery from sequences detected in prospective clinical samples. Genome recovery was defined as the percentage of SARS-CoV-2 genome nucleotide sites with greater than 10X coverage for Illumina and 20X coverage for Nanopore.

### Comparison between diagnostic assays

Samples were initially tested by one of twelve clinical diagnostic assays; samples positive for SARS-CoV-2 were then subjected to the ARTIC sequencing process. Thus, the sensitivities of the different diagnostic platforms can be compared based on final genomic data. The comparative measure we used, and is described here, represents the number of samples that were deemed positive by a diagnostics platform, but were subsequently unsupported by the sequencing data.

This screening identified 1291, 972, 899 and 441 mapped read sets within the NORW, EDIN, PORT and NORT datasets, respectively, from a total of 3,603 samples. Samples where no diagnostic platform were filtered out, giving a final count of 3,107. The number of samples evaluated by each diagnostic platform varied (Table 3, Figure 3).

**Table 3:** Summary of samples evaluated using each diagnostic platform and deemed positive for SARS-CoV-2 and the number and proportion that were subsequently unsupported by sequencing data.

| Diagnostic Platform | Samples negative by sequencing (%) | Total number of samples evaluated | ARTIC sensitivity |
| --- | --- | --- | --- |
| Roche Lightcycler 480 II | 23 (3.7%) | 626 | 96.3% |
| AusDiagnostics | 48 (8.4%) | 562 | 91.5% |
| Roche Cobas | 13 (2.8%) | 467 | 97.2% |
| Hologic Panther | 34 (11.6%) | 293 | 88.4% |
| Corman RdRp | 1 (0.4%) | 245 | 99.6% |
| Roche Flow | 3 (1.1%) | 273 | 98.9% |
| ABI QuantStudio 5 | 10 (4.4%) | 228 | 95.6% |
| Seegene Nimbus | 11 (5.0%) | 219 | 95.0% |
| Qiagen Rotorgene* | 6 (7.5%) | 80 | - |
| Abbott M2000* | 6 (11.1%) | 54 | - |
| Thermo TaqPath* | 3 (6.1%) | 49 | - |
| Xpert* | 0 (0%) | 11 | - |

\*Total number of samples was insufficient to reliably determine the sensitivity of ARTIC.

**Figure 3:**
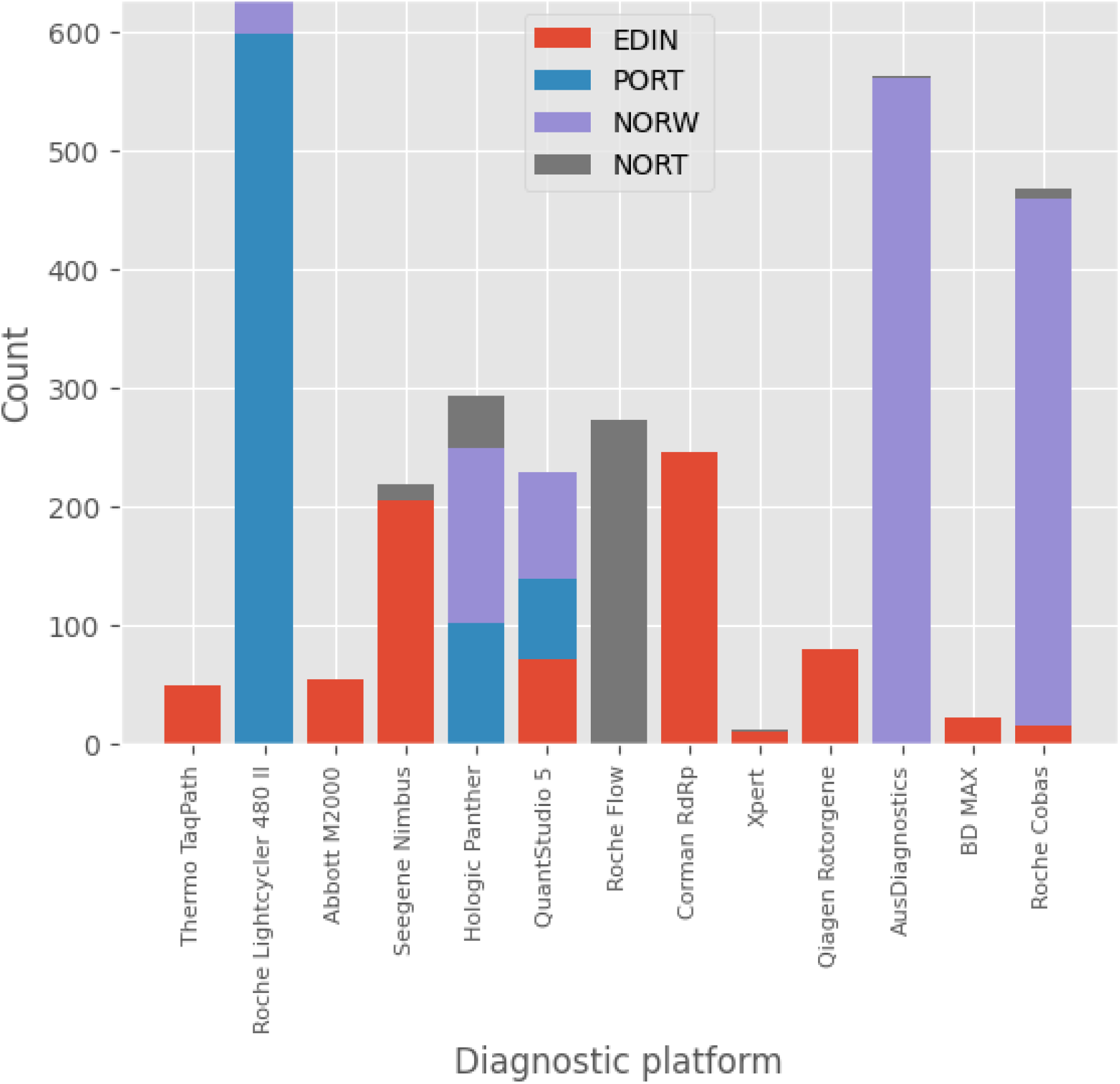
SARS-CoV-2 positive samples included in this study. The chart was broken down by diagnostic platform used (y-axis) and sample site (NORW, PORT, NORT, EDIN) (coloured according to the key).

Sequenced reads from samples were mapped to the Wuhan-Hu-1 SARS-CoV-2 reference genome (MN908947.3). Samples were considered ARTIC negative if they failed to meet a minimum threshold of: 2X total genome coverage; or 2 times the genome recovery of the negative control. Illumina data required additional criteria to be met, specifically: the total number of fully-mapped reads had to be both 5X greater than the negative control and greater than an absolute value of 30. These criteria identified a number of unsupported samples, the number of which varied between each of the platforms surveyed (Figure 4A).

**Figure 4:**
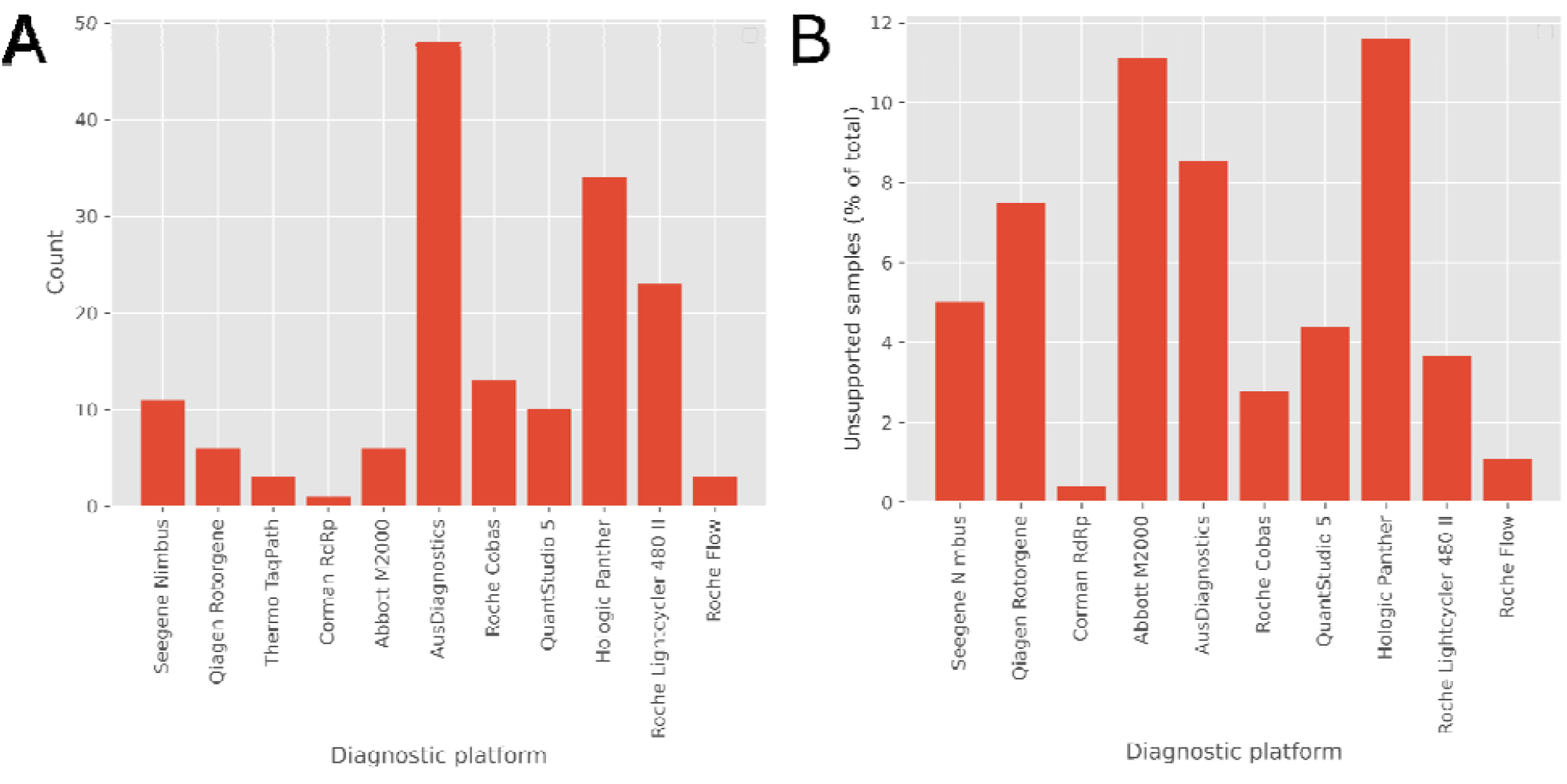
Proportion of discrepancies between diagnostic platforms and sequencing content. (A) absolute count of samples where diagnostic platform (y-axis) reported a positive but ARTIC was negative (B) proportion of false negative samples per platform. Only platforms with >50 samples are shown.

Only Hologic Panther, Ausdiagnostics, ABI QuantStudio, Abbott M200, Seegene Nimbus and Roche platforms provided a sufficient number of true positive samples (>50) to allow for meaningful comparison. The sensitivity of ARTIC on positive samples from the various platforms is listed in Table 3. The number and proportion of samples negative by ARTIC for each platform is shown in Figure 4.

We also compared genome recovery rate of the different platforms within acceptable Ct values (less than Ct 31) (Figure 5). Genome recovery was above average for Ausdiagnostics, QuantStudio 5, Seegene Nimbus, and Roche platforms when compared to all other platforms..

**Figure 5:**
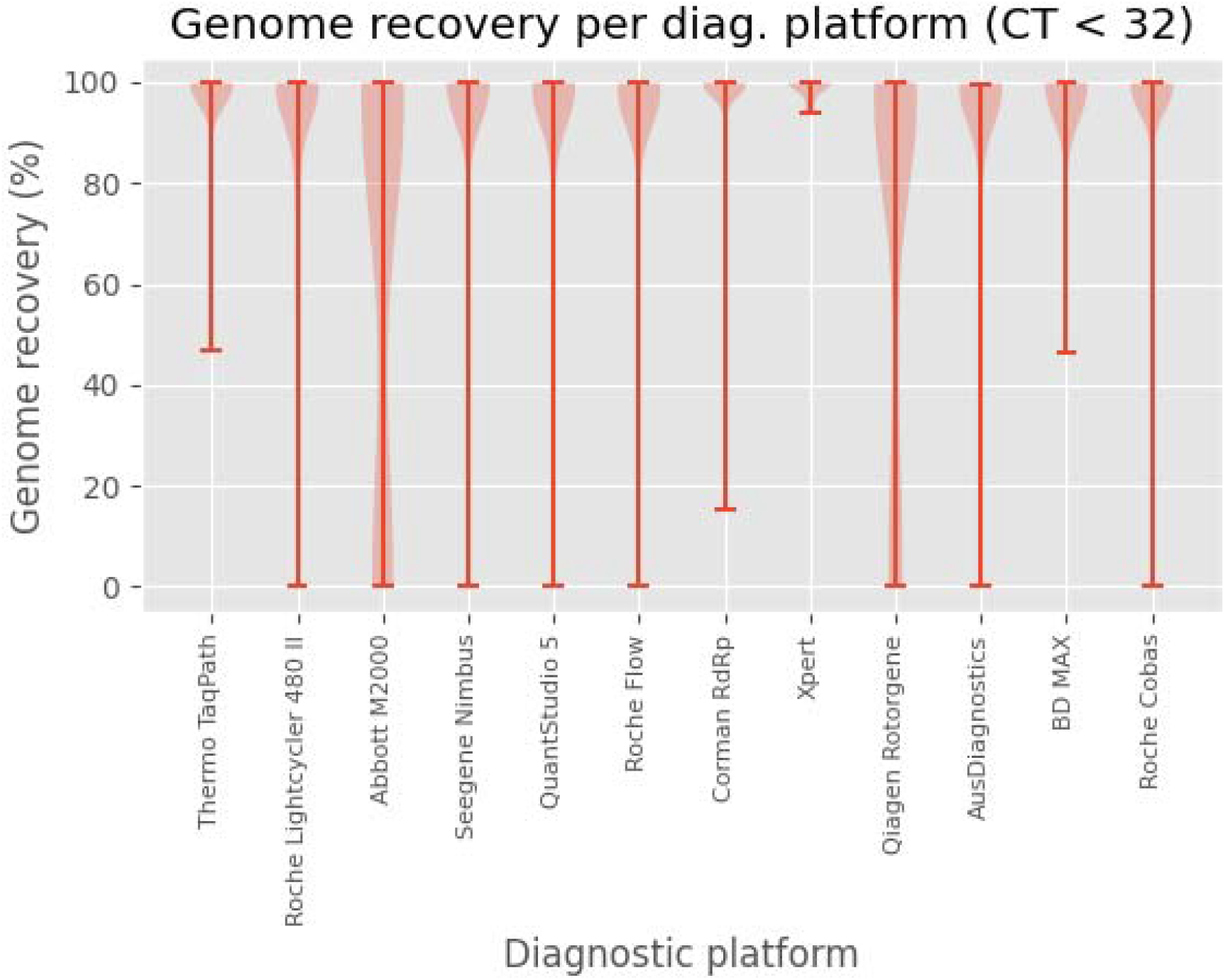
Genome recovery per diagnostic platforms for samples. Includes samples for 13 diagnostic platforms under Ct 32. Hologic Panther was not included as it does not use RT-qPCR and does not report a Ct value.

## Discussion

ARTIC based whole genome sequencing is used to investigate the genomic epidemiology of SARS-CoV-2 and to identify novel and existing variants under investigation (VUIs) and variants of concern (VOCs). ARTIC isn’t typically used as a diagnostic test, it is applied to SARS-CoV-2 samples determined positive by one of many available nucleic acid amplification tests (RT-qPCR, LAMP etc). Diagnostic tests for SARS-CoV-2 have different reported sensitivities and thresholds for determining whether a sample is positive or not. It is thus important to understand the limitations of the ARTIC protocol and how sequencing data compares with the results provided by diagnostic platforms when both are applied to the same samples. Here we define the analytical and clinical sensitivity of ARTIC for the detection of SARS-CoV-2 in mock and clinical samples respectively. We also define the RT-qPCR Ct thresholds that provide high quality sequencing results (>80% and 90% genome recovery) so that only positive samples likely to provide useful information can be sequenced.

By processing SARS-CoV-2 samples of known concentrations, in triplicate, we established the LoD for the ARTIC protocol as 25-50 viral copies per mL (Figure 1). These values can be compared with listed sensitivities of diagnostic tests, and showed ARTIC was comparable, if not slightly more sensitive than the platforms described (Table 1). However, genome recovery (% of genome covered at >10x) at the LoD is <10% (Fig 1A) and unlikely to provide useful lineage information. Genome recovery increases to an average of 35% when 100 viral copies are present in the ARTIC PCR which equates to approx Ct 31 (for a sensitive RT-qPCR assay). Therefore it could be estimated that positive clinical samples with Ct <31 would be required to provide >50% genome recovery using ARTIC, assuming the diagnostic assays are very sensitive and the volume of RNA extract tested in the RT-qPCR is similar to ARTIC (11ul).

Sequenced reads from known concentrations of SARS-CoV-2 allowed us to define the metrics and thresholds to determine whether a sample was positive for SARS-CoV-2. The clearest metric was ‘genome recovery’, defined as the number of SARS-CoV-2 genome bases that had a genome coverage that exceeded a defined threshold (10X for Illumina, 20X for Nanopore)(Figure 1A). Other useful metrics were mean genome coverage and total number of mapped reads when mapped to the SARS-CoV-2 reference genome (Figure 1B). Additional metrics were included to account for barcode crosstalk and laboratory/reagent contamination. These metrics and thresholds were then used in the RonaLDO package.

The RonaLDO package includes modules for calculating the metrics and assessing them with defined thresholds; and a plotting module for generating charts for reporting purposes. Default thresholds are in line with the data observed here, but are easily modified by specifying command line parameters. RonaLDO was developed as an open source package to encourage the community to apply it to their own data, allowing for further refinement of the metrics and reporting tools.

When RonaLDO was run on the real-world dataset in this paper from COG-UK, it is clear that the results seen in the mock samples mirror real clinical samples (Figure 2). Positive samples with Ct values <32 resulted in high genome recovery (>80% on average). This is slightly higher than expected based on the mock community analysis, but this is likely explained by some tests having lower sensitivity (hence Ct 31 =>100 viral copies).

ARTIC demonstrated high sensitivity (>88%) compared to the various diagnostics tests (Table 3, Figure 4). The AusDx and Hologic and Abbott M2000 (laboratory developed test) were the assays with the highest false negative ARTIC results, suggesting these assays are the most sensitive, or potentially produce more false positive results. AusDx utilises nested PCR and Hologic is TMA based which could explain improved sensitivity for these tests. The manual RDRP assay (and the Xpert test, but very low numbers were tested) had the lowest false negative ARTIC rate at 0.4% -this would suggest that it wasn’t as sensitive with higher Ct values containing more viral genome copies than expected. When comparing diagnostic tests close to the LoD, discrepancies are expected due to the low number of viral genome copies in the samples and the chance of testing an aliquot without any target present. Some of this variation can also be attributed to different sample handling processes, such as re-extraction of RNA from primary sample using a different RNA extraction method a number of days after the original test was run (Hologic and Roche Cobas).

Routine use of ARTIC on samples ascribed as negative by diagnostic platforms but accidentally sent for sequencing confirmed them to be negative. This supports the premise that sequencing data can support diagnostic tests.

Focussing on genome recovery, the important metric for genome sequencing success, we compared all the RT-qPCR based diagnostics tests on samples with Ct <32. As you can see in Figure 5, the vast majority of samples for most platforms had ARTIC genome recovery >80%. The exceptions were the Abbott M2000 and Qiagen Rotorgene laboratory developed tests which have a wider range of genome recovery percentages and a significant number of samples with very low (or no) genome recovery. This suggests the potential of false positive results produced by these tests.

It is clear that the degree of variation in diagnostics assays represents a logistical problem if positive samples are subjected to ARTIC and subsequently found to be negative or have low genome recovery (thereby wasting sequencing effort and money). For those wishing to maximise success of ARTIC for phylogenetic analysis, we recommend that samples should be less than Ct 31. Where the diagnostic test does not provide a quantifiable Ct value, we recommend an RT-qPCR quantification step be added to the sequencing protocol.

This study suffers from a number of limitations. The focus of our study has been on the implications of variation in the outcome of diagnostic platforms themselves on subsequent sequencing success. However, we acknowledge that the RNA extraction method may play a yet unknown role in this variation. As samples presented here were always extracted from primary material, it was not possible to test the effects of sequencing material that had been subjected to different preparatory processes, for instance, comparing RNA extracted from lysate produced from some diagnostic tests versus RNA extracted from the primary material. It should be noted that this comparison was limited only to the diagnostics assays used in the laboratories working directly with the NORW, PORT, EDIN and NORT COG-UK regions.

## Conclusions

In conclusion, our data demonstrates that amplicon sequencing using the ARTIC protocol is highly sensitive for the detection of SARS-CoV-2 and can assist diagnostics tests. In a controlled experiment we demonstrated that ARTIC had a LoD between 25 - 50 virus particles per mL. When we compared sequencing data with the results of SARS-CoV-2 diagnostic platforms we found that positives detected by diagnostic platforms were generally supported by sequencing data. There were discrepancies for all platforms, but platforms that used RT-qPCR provided the best predictor that the sample would sequence successfully. The data here provides no judgement on which protocol is more appropriate for SARS-CoV-2 testing, rather that a positive diagnostics assay result does not guarantee that the same sample will be sequenced successfully. For those wishing to maximise the success of sample sequencing for phylogenetic analysis, we recommend that samples should be less than Ct 31. When sequencing samples for which the diagnostic test does not provide a quantifiable Ct value, we recommend an explicit quantification step be added to the sequencing protocol.

## Data Availability

Raw sequence data from clinical samples were deposited in and are available from the European Nucleotide Archive under BioProject accession number PRJEB37886. All consensus genomes are available from COG-UK (https://www.cogconsortium.uk) and high-quality genomes are also available from GISAID. Sequence data from LoD experiments are available under BioProject accession number PRJEB41469.

## Ethical approval

The COVID-19 Genomics UK Consortium has been given approval by Public Health England’s Research Ethics and Governance Group (PHE R&D Ref: NR0195).

## Acknowledgements

The authors would like to thank COG-UK Consortium Study Group for their contributions. The authors would like to thank Carol Davies-Sala for useful input on the manuscript. The authors would like to thank Judith Pell for enhancing the manuscript. The authors would also like to thank Angela H. Beckett, Kate F. Cook, Christopher Fearn and Salman Goudarzi from the University of Portsmouth and Sharon Glaysher Portsmouth Hospitals University NHS Trust for their work in generating the sequencing data. The authors would also like to thank Allyson Lloyd, Sarah Wyllie, Scott Elliott, Kelly Bicknell and Robert Impey from Portsmouth Hospitals University NHS Trust; Mohammed O. Hassan-Ibrahim, Cassandra S. Malone, Benjamin J. Cogger, Lisa J. Easton, Nicola J. Chaloner and Thomas Somas from Brighton and Sussex University Hospitals NHS Trust; Kordo Saeed, Eleri Wilson-Davies, Emanuela Pelosi, Jacqui A Prieto, Jacqui A Prieto, Adhyana I. K. Mahanama, Buddhini Samaraweera, Siona Silviera and Sarah Jeremiah from University Hospital Southampton NHS Foundation Trust for providing samples for sequencing. The authors acknowledge the contributions of Helen Wheeler, Matthew Harvey, Thea Sass and Helen Umpleby to this study.

## Author contributions

All authors have read this manuscript and consented to its publication. NA wrote bioinformatics software for analysis and analysed the data. AJT and SD performed ARTIC LOD experiments. NA and AJP wrote the first draft of the manuscript. NA, NJL, AJP, AJT, GLK, JOG, MB and SRobson revised the manuscript. MB, ML, SRobson, ACD, SN, MMcH and SRooke ran analyses for their respective sites and contributed data. SD contributed NORW samples and associated data. JQ and JOG conceived the study and provided overall leadership.

## References

1. Huang C, Wang Y, Li X, Ren L, Zhao J, Hu Y, et al. Clinical features of patients infected with 2019 novel coronavirus in Wuhan, China. The Lancet. 2020 Feb 15;395(10223):497–506.

2. Dong E, Du H, Gardner L. An interactive web-based dashboard to track COVID-19 in real time. The Lancet Infectious Diseases. 2020 May 1;20(5):533–4.

3. Quick J, Grubaugh ND, Pullan ST, Claro IM, Smith AD, Gangavarapu K, et al. Multiplex PCR method for MinION and Illumina sequencing of Zika and other virus genomes directly from clinical samples. Nature protocols. 2017;12(6):1261.

4. Artic Network [Internet]. [cited 2020 Jul 6]. Available from: https://artic.network/

5. Shu Y, McCauley J. GISAID: Global initiative on sharing all influenza data – from vision to reality. Eurosurveillance. 2017 Mar 30;22(13):30494.

6. SARS-CoV-2 Q Control 01 [Internet]. Qnostics; [cited 2020 Oct 30]. Available from: https://www.qnostics.com/wp-content/uploads/2020/05/SCV2QC01-A-RBPL3169-Rev01.pdf

7. Baker DJ, Kay GL, Aydin A, Le-Viet T, Rudder S, Tedim AP, et al. CoronaHiT: large scale multiplexing of SARS-CoV-2 genomes using Nanopore sequencing. bioRxiv. 2020 Jun 24;2020.06.24.162156.

8. Quick J. nCoV-2019 sequencing protocol v2. 2020 Apr 9 [cited 2020 Apr 25]; Available from: https://www.protocols.io/view/ncov-2019-sequencing-protocol-v2-bdp7i5rn

9. ncov2019-artic-nf [Internet]. Available from: https://github.com/connor-lab/ncov2019-artic-nf

10. Bashton M. articSlurmy [Internet]. Available from: https://github.com/MattBashton/articSlurmy

11. Alikhan N-F. ronaLDO [Internet]. Available from: https://github.com/happykhan/ronaldo

12. Adaptation of Connor Lab Nextflow pipeline for running the ARTIC network’s field bioinformatics tools [Internet]. [cited 2020 Jul 6]. Available from: https://github.com/quadram-institute-bioscience/ncov2019-artic-nf

13. Smith E, Zhen W, Manji R, Schron D, Duong S, Berry GJ. Analytical and Clinical Comparison of Three Nucleic Acid Amplification Tests for SARS-CoV-2 Detection. McAdam AJ, editor. J Clin Microbiol. 2020 Aug 24;58(9):e01134–20.

14. AusDiagnostics. SARS-COV-2, INFLUENZA AND RSV 8-WELL [Internet]. 2020 Apr p. 20081-r01.1. Report No.: 20081. Available from: https://www.ausdx.com/qilan/Products/20081-r01.1.pdf

15. Xpert® Xpress SARS-CoV-2 [Internet]. Cepheid; Available from: https://www.cepheid.com/coronavirus-product-resources

17. Qualitative assay for use on the cobas® 6800/8800 Systems [Internet]. [cited 2020 Jul 6]. Available from: https://www.who.int/diagnostics_laboratory/eul_0504-046-00_cobas_sars_cov2_qualitative_assay_ifu.pdf

18. Page AJ, Mather AE, Le Viet T, Meader EJ, Alikhan N-FJ, Kay GL, et al. Large scale sequencing of SARS-CoV-2 genomes from one region allows detailed epidemiology and enables local outbreak management. medRxiv. 2020 Jan 1;2020.09.28.20201475.

