## Supplementary Material 1 for "Defining the analytical and clinical sensitivity of the ARTIC method for the detection of SARS-CoV-2"

**Funding acquisition,** **Leadership and supervision, Metadata curation,** **Project administration,** **Samples and logistics,** **Sequencing and analysis,** **Software and analysis tools, and Visualisation:**

Dr Samuel C Robson PhD ^13^.

**Funding acquisition,** **Leadership and supervision, Metadata curation,** **Project administration,** **Samples and logistics,** **Sequencing and analysis, and Software and analysis tools:**

Prof Nicholas J Loman PhD ^41^ and Dr Thomas R Connor PhD ^10,^ ^69^.

**Leadership and supervision, Metadata curation,** **Project administration,** **Samples and logistics,** **Sequencing and analysis,** **Software and analysis tools, and Visualisation:**

Dr Tanya Golubchik PhD ^5^.

**Funding acquisition,** **Metadata curation,** **Samples and logistics,** **Sequencing and analysis,** **Software and analysis tools, and Visualisation:**

Dr Rocio T Martinez Nunez PhD ^42^.

**Funding acquisition,** **Leadership and supervision, Metadata curation,** **Project administration, and Samples and logistics:**

Dr Catherine Ludden PhD ^88^.

**Funding acquisition,** **Leadership and supervision, Metadata curation,** **Samples and logistics, and Sequencing and analysis:**

Dr Sally Corden PhD ^69^.

**Funding acquisition,** **Leadership and supervision, Project administration,** **Samples and logistics, and Sequencing and analysis:**

Ian Johnston ^99^ and Dr David Bonsall PhD ^5^.

**Funding acquisition,** **Leadership and supervision, Sequencing and analysis,** **Software and analysis tools, and Visualisation:**

Prof Colin P Smith PhD ^87^ and Dr Ali R Awan PhD ^28^.

**Funding acquisition,** **Samples and logistics,** **Sequencing and analysis,** **Software and analysis tools, and Visualisation:**

Dr Giselda Bucca PhD ^87^.

**Leadership and supervision, Metadata curation,** **Project administration,** **Samples and logistics, and Sequencing and analysis:**

Dr M. Estee Torok FRCP ^22,^ ^101^.

**Leadership and supervision, Metadata curation,** **Project administration,** **Samples and logistics, and Visualisation:**

Dr Kordo Saeed MD/ FRCPath ^81,^ ^110^ and Dr Jacqui A Prieto PhD ^83,^ ^109^.

**Leadership and supervision, Metadata curation,** **Project administration,** **Sequencing and analysis, and Software and analysis tools:**

Dr David K Jackson PhD ^99^.

**Metadata curation,** **Project administration,** **Samples and logistics,** **Sequencing and analysis, and Software and analysis tools:**

Dr William L Hamilton PhD ^22^.

**Metadata curation,** **Project administration,** **Samples and logistics,** **Sequencing and analysis, and Visualisation:**

Dr Luke B Snell MSc/ MBBS ^11^.

**Funding acquisition,** **Leadership and supervision, Metadata curation, and Samples and logistics:**

Dr Catherine Moore ^69^.

**Funding acquisition,** **Leadership and supervision, Project administration, and Samples and logistics:**

Dr Ewan M Harrison PhD ^99,^ ^88^.

**Leadership and supervision, Metadata curation,** **Project administration, and Samples and logistics:**

Dr Sonia Goncalves PhD ^99^ and Dr Leigh M Jackson Ph.D. ^91^.

**Leadership and supervision, Metadata curation,** **Samples and logistics, and Sequencing and analysis:**

Prof Ian G Goodfellow PhD ^24^, Dr Derek J Fairley PhD ^3,^ ^72^, Prof Matthew W Loose PhD ^18^ and Joanne Watkins MSc ^69^.

**Leadership and supervision, Metadata curation,** **Samples and logistics, and Software and analysis tools:**

Rich Livett MSc ^99^.

**Leadership and supervision, Metadata curation,** **Samples and logistics, and Visualisation:**

Dr Samuel Moses MD ^25,^ ^106^.

**Leadership and supervision, Metadata curation,** **Sequencing and analysis, and Software and analysis tools:**

Dr Roberto Amato PhD ^99^, Dr Sam Nicholls PhD ^41^ and Dr Matthew Bull PhD ^69^.

**Leadership and supervision, Project administration,** **Samples and logistics, and Sequencing and analysis:**

Prof Darren L Smith PhD ^37,^ ^58,^ ^105^.

**Leadership and supervision, Sequencing and analysis,** **Software and analysis tools, and Visualisation:**

Dr Jeff Barrett PhD ^99^ and Prof David M Aanensen PhD ^14,^ ^114^.

**Metadata curation,** **Project administration,** **Samples and logistics, and Sequencing and analysis:**

Dr Martin D Curran PhD ^65^, Dr Surendra Parmar PhD ^65^, Dr Dinesh Aggarwal MRCP ^95,^ ^99,^ ^64^ and Dr James G Shepherd MBChB/MRCP ^48^.

**Metadata curation,** **Project administration,** **Sequencing and analysis, and Software and analysis tools:**

Dr Matthew D Parker PhD ^93^.

**Metadata curation,** **Samples and logistics,** **Sequencing and analysis, and Visualisation:**

Dr Sharon Glaysher PhD ^61^.

**Metadata curation,** **Sequencing and analysis,** **Software and analysis tools, and Visualisation:**

Dr Matthew Bashton PhD ^37,^ ^58^, Dr Anthony P Underwood PhD ^14,^ ^114^, Dr Nicole Pacchiarini PhD ^69^ and Dr Katie F Loveson PhD ^77^.

**Project administration,** **Sequencing and analysis,** **Software and analysis tools, and Visualisation:**

Dr Alessandro M Carabelli PhD ^88^.

**Funding acquisition,** **Leadership and supervision, and Metadata curation:**

Dr Kate E Templeton PhD ^53,^ ^90^.

**Funding acquisition,** **Leadership and supervision, and Project administration:**

Dr Cordelia F Langford PhD ^99^, John Sillitoe BEng ^99^, Dr Thushan I de Silva PhD ^93^ and Dr Dennis Wang PhD ^93^.

**Funding acquisition,** **Leadership and supervision, and Sequencing and analysis:**

Prof Dominic Kwiatkowski ^99,^ ^107^, Prof Andrew Rambaut DPhil ^90^, Dr Justin O’Grady PhD ^70,^ ^89^ and Dr Simon Cottrell PhD ^69^.

**Leadership and supervision, Metadata curation, and Sequencing and analysis:**

Prof Matthew T.G. Holden PhD ^68^ and Prof Emma C Thomson PhD/FRCP ^48^.

**Leadership and supervision, Project administration, and Samples and logistics:**

Dr Husam Osman PhD ^64,^ ^36^, Dr Monique Andersson PhD ^59^, Prof Anoop J Chauhan ^61^ and Dr Mohammed O Hassan-Ibrahim PhD/FRCPath ^6^.

**Leadership and supervision, Project administration, and Sequencing and analysis:**

Dr Mara Lawniczak ^99^.

**Leadership and supervision, Samples and logistics, and Sequencing and analysis:**

Prof Ravi Kumar Gupta PhD ^88,^ ^113^, Dr Alex Alderton PhD ^99^, Dr Meera Chand ^66^, Dr Chrystala Constantinidou PhD ^94^, Dr Meera Unnikrishnan PhD ^94^, Prof Alistair C Darby PhD ^92^, Prof Julian A Hiscox PhD ^92^ and Prof Steve Paterson PhD ^92^.

**Leadership and supervision, Sequencing and analysis, and Software and analysis tools:**

Dr Inigo Martincorena ^99^, Prof David L Robertson PhD ^48^, Dr Erik M Volz PhD ^39^, Dr Andrew J Page PhD ^7^ and Prof Oliver G Pybus DPhil ^23^.

**Leadership and supervision, Sequencing and analysis, and Visualisation:**

Dr Andrew R Bassett PhD ^99^.

**Metadata curation,** **Project administration, and Samples and logistics:**

Dr Cristina V Ariani PhD ^99^, Dr Michael H Spencer Chapman MBBS ^99,^ ^88^, Dr Kathy K Li MBBCh/FRCPath ^48^, Dr Rajiv N Shah BMBS/MRCP/MSc ^48^, Dr Natasha G Jesudason MBChB MRCP FRCPath ^48^ and Dr Yusri Taha MD/PhD ^50^.

**Metadata curation,** **Project administration, and Sequencing and analysis:**

Martin P McHugh MSc ^53^, Dr Rebecca Dewar PhD ^53^.

**Metadata curation,** **Samples and logistics, and Sequencing and analysis:**

Dr Aminu S Jahun PhD ^24^, Dr Claire McMurray PhD ^41^, Ms Sarojini Pandey MSc ^84^, Dr James P McKenna PhD ^3^, Dr Andrew Nelson PhD ^58,^ ^105^, Dr Gregory R Young PhD ^37,^ ^58^, Dr Clare M McCann PhD ^58,^ ^105^ and Mr Scott Elliott ^61^.

**Metadata curation,** **Samples and logistics, and Visualisation:**

Ms Hannah Lowe MSc ^25^.

**Metadata curation,** **Sequencing and analysis, and Software and analysis tools:**

Dr Ben Temperton Ph.D. ^91^, Dr Sunando Roy PhD ^82^, Dr Anna Price PhD ^10^, Dr Sara Rey PhD ^69^ and Mr Matthew Wyles ^93^.

**Metadata curation,** **Sequencing and analysis, and Visualisation:**

Stefan Rooke MSc ^90^ and Dr Sharif Shaaban PhD ^68^.

**Project administration,** **Samples and logistics,** **Sequencing and analysis:**

Dr Mariateresa de Cesare PhD ^98^.

**Project administration,** **Samples and logistics, and Software and analysis tools:**

Laura Letchford BSc ^99^.

**Project administration,** **Samples and logistics, and Visualisation:**

Miss Siona Silveira MSc ^81^, Dr Emanuela Pelosi FRCPath ^81^ and Dr Eleri Wilson-Davies MD/FRCPath ^81^.

**Samples and logistics,** **Sequencing and analysis, and Software and analysis tools:**

Dr Myra Hosmillo PhD ^24^.

**Sequencing and analysis,** **Software and analysis tools, and Visualisation:**

Áine O'Toole MSc ^90^, Dr Andrew R Hesketh PhD ^87^, Mr Richard Stark MSc ^94^, Dr Louis du Plessis PhD ^23^, Dr Chris Ruis PhD ^88^, Dr Helen Adams PhD ^4^ and Dr Yann Bourgeois PhD ^76^.

**Funding acquisition, and Leadership and supervision:**

Dr Stephen L Michell PhD ^91^, Prof Dimitris Grammatopoulos PhD/FRCPath ^84,^ ^112^, Dr Jonathan Edgeworth PhD/FRCPath ^12^, Prof Judith Breuer MD ^30,^ ^82^, Prof John A Todd PhD ^98^ and Dr Christophe Fraser PhD ^5^.

**Funding acquisition, and Project administration:**

Dr David Buck PhD ^98^ and Michaela John BSc ^9^.

**Leadership and supervision, and Metadata curation:**

Dr Gemma L Kay PhD ^70^.

**Leadership and supervision, and Project administration:**

Steve Palmer ^99^, Prof Sharon J Peacock ^88,^ ^64^ and David Heyburn ^69^.

**Leadership and supervision, and Samples and logistics:**

Danni Weldon BSc ^99^, Dr Esther Robinson PhD ^64,^ ^36^, Prof Alan McNally PhD ^41,^ ^86^, Dr Peter Muir PhD ^64^, Dr Ian B Vipond PhD ^64^, Dr John BoYes MBChB ^29^, Dr Venkat Sivaprakasam PhD ^46^, Dr Tranprit Saluja FRCPath/MD ^75^, Dr Samir Dervisevic FRCPath ^54^ and Dr Emma J Meader FRCPath ^54^.

**Leadership and supervision, and Sequencing and analysis:**

Dr Naomi R Park PhD ^99^, Karen Oliver BSc ^99^, Dr Aaron R Jeffries Ph.D. ^91^, Dr Sascha Ott PhD ^94^, Dr Ana da Silva Filipe PhD ^48^, Dr David A Simpson PhD ^72^ and Dr Chris Williams MB BS ^69^.

**Leadership and supervision, and Visualisation:**

Dr Jane AH Masoli MBChB ^73,^ ^91^.

**Metadata curation, and Samples and logistics:**

Dr Bridget A Knight PhD. ^73,^ ^91^, Dr Christopher R Jones Ph.D. ^73,^ ^91^, Mr Cherian Koshy MSc CSci FIBMS ^1^, Miss Amy Ash BSc ^1^, Dr Anna Casey PhD ^71^, Dr Andrew Bosworth PhD ^64,^ ^36^, Dr Liz Ratcliffe PhD ^71^, Dr Li Xu-McCrae PhD ^36^, Miss Hannah M Pymont MSc ^64^, Ms Stephanie Hutchings ^64^, Dr Lisa Berry PhD ^84^, Ms Katie Jones MSc ^84^, Dr Fenella Halstead PhD ^46^, Mr Thomas Davis MSc ^21^, Dr Christopher Holmes PhD ^16^, Prof Miren Iturriza-Gomara PhD ^92^, Dr Anita O Lucaci PhD ^92^, Dr Paul Anthony Randell MBBCh ^38,^ ^104^, Dr Alison Cox PhD ^38,^ ^104^, Pinglawathee Madona ^38,^ ^104^, Dr Kathryn Ann Harris PhD ^30^, Dr Julianne Rose Brown PhD ^30^, Dr Tabitha W Mahungu FRCPath ^74^, Dr Dianne Irish-Tavares FRCPath ^74^, Dr Tanzina Haque FRCPath PhD ^74^, Dr Jennifer Hart MRCP ^74^, Mr Eric Witele MSc ^74^, Mrs Melisa Louise Fenton DipHE ^75^, Mr Steven Liggett ^79^, Dr Clive Graham MD ^56^, Ms Emma Swindells Bsc ^57^, Ms Jennifer Collins BSc ^50^, Mr Gary Eltringham BSc ^50^, Ms Sharon Campbell MSc ^17^, Dr Patrick C McClure PhD ^97^, Dr Gemma Clark PhD ^15^, Dr Tim J Sloan PhD ^60^, Mr Carl Jones ^15^ and Dr Jessica Lynch PhD MBChB ^2,^ ^111^.

**Metadata curation, and Sequencing and analysis:**

Dr Ben Warne MRCP ^8^, Steven Leonard PhD ^99^, Jillian Durham BSc ^99^, Dr Thomas Williams MD ^90^, Dr Sam T Haldenby PhD ^92^, Dr Nathaniel Storey PhD ^30^, Dr Nabil-Fareed Alikhan PhD ^70^, Dr Nadine Holmes PhD ^18^, Dr Christopher Moore PhD ^18^, Mr Matthew Carlile BSc ^18^, Malorie Perry MSc ^69^, Dr Noel Craine DPhil ^69^, Prof Ronan A Lyons MD ^80^, Miss Angela H Beckett MSc ^13^, Salman Goudarzi PhD ^77^, Christopher Fearn MRes ^77^, Kate Cook ^77^, Hannah Dent BSc ^77^ and Hannah Paul MRes ^77^.

**Metadata curation, and Software and analysis tools:**

Robert Davies ^99^.

**Project administration, and Samples and logistics:**

Beth Blane BSc ^88^, Sophia T Girgis MSc ^88^, Dr Mathew A Beale PhD ^99^, Katherine L Bellis ^99,^ ^88^, Matthew J Dorman ^99^, Eleanor Drury ^99^, Leanne Kane ^99^, Sally Kay ^99^, Dr Samantha McGuigan ^99^, Dr Rachel Nelson PhD ^99^, Liam Prestwood ^99^, Dr Shavanthi Rajatileka PhD ^99^, Dr Rahul Batra MD ^12^, Dr Rachel J Williams PhD ^82^, Dr Mark Kristiansen PhD ^82^, Dr Angie Green PhD ^98^, Miss Anita Justice MSc ^59^, Dr Adhyana I.K Mahanama MD ^81,^ ^102^ and Dr Buddhini Samaraweera MD ^81,^ ^102^.

**Project administration, and Sequencing and analysis:**

Dr Nazreen F Hadjirin PhD ^88^ and Dr Joshua Quick PhD ^41^.

**Project administration, and Software and analysis tools:**

Mr Radoslaw Poplawski BSc ^41^.

**Samples and logistics, and Sequencing and analysis:**

Leanne M Kermack MSc ^88^, Nicola Reynolds PhD ^7^, Grant Hall BS ^24^, Yasmin Chaudhry BSc ^24^, Malte L Pinckert MPhil ^24^, Dr Iliana Georgana PhD ^24^, Dr Robin J Moll PhD ^99^, Dr Alicia Thornton ^66^, Dr Richard Myers ^66^, Dr Joanne Stockton PhD ^41^, Miss Charlotte A Williams BSc ^82^, Dr Wen C Yew PhD ^58^, Alexander J Trotter MRes ^70^, Miss Amy Trebes MSc ^98^, Mr George MacIntyre-Cockett BSc ^98^, Alec Birchley MSc ^69^, Alexander Adams BSc ^69^, Amy Plimmer ^69^, Bree Gatica-Wilcox MPhil ^69^, Dr Caoimhe McKerr PhD ^69^, Ember Hilvers MA ^69^, Hannah Jones ^69^, Dr Hibo Asad PhD ^69^, Jason Coombes BSc ^69^, Johnathan M Evans MSc ^69^, Laia Fina ^69^, Lauren Gilbert A-Levels ^69^, Lee Graham BSc ^69^, Michelle Cronin ^69^, Sara Kumziene-SummerhaYes MSc ^69^, Sarah Taylor ^69^, Sophie Jones MSc ^69^, Miss Danielle C Groves BA ^93^, Mrs Peijun Zhang MSc ^93^, Miss Marta Gallis MSc ^93^ and Miss Stavroula F Louka MSc ^93^.

**Samples and logistics, and Software and analysis tools:**

Dr Igor Starinskij Msc MRCP ^48^.

**Sequencing and analysis, and Software and analysis tools:**

Dr Chris J Illingworth PhD ^47^, Dr Chris Jackson PhD ^47^, Ms Marina Gourtovaia MSc ^99^, Gerry Tonkin-Hill ^99^, Kevin Lewis ^99^, Dr Jaime M Tovar-Corona PhD ^99^, Dr Keith James PhD ^99^, Dr Laura Baxter PhD ^94^, Dr Mohammad T. Alam PhD ^94^, Dr Richard J Orton PhD ^48^, Dr Joseph Hughes PhD ^48^, Dr Sreenu Vattipally PhD ^48^, Dr Manon Ragonnet-Cronin PhD ^39^, Dr Fabricia F. Nascimento PhD ^39^, Mr David Jorgensen MSc ^39^, Ms Olivia Boyd MSc ^39^, Ms Lily Geidelberg MSc ^39^, Dr Alex E Zarebski PhD ^23^, Dr Jayna Raghwani PhD ^23^, Dr Moritz UG Kraemer DPhil ^23^, Joel Southgate MSc ^10,^ ^69^, Dr Benjamin B Lindsey MRCP ^93^ and Mr Timothy M Freeman MPhil ^93^.

**Software and analysis tools, and Visualisation:**

Jon-Paul Keatley ^99^, Dr Joshua B Singer PhD ^48^, Leonardo de Oliveira Martins PhD ^70^, Dr Corin A Yeats PhD ^14^, Dr Khalil Abudahab PhD ^14,^ ^114^, Mr Ben EW Taylor MEng ^14,^ ^114^ and Mirko Menegazzo ^14^.

**Leadership and supervision:**

Prof John Danesh ^99^, Wendy Hogsden MSc ^46^, Dr Sahar Eldirdiri MBBS MSc FRCPath ^21^, Mrs Anita Kenyon MSc ^21^, Dr Jenifer Mason MBBS ^43^, Mr Trevor I Robinson MSc ^43^, Prof Alison Holmes MD ^38,^ ^103^, Dr James Price PhD ^38,^ ^103^, Prof John A Hartley PhD ^82^, Dr Tanya Curran PhD ^3^, Dr Alison E Mather PhD ^70^, Dr Giri Shankar ^69^, Dr Rachel Jones ^69^, Dr Robin Howe ^69^ and Dr Sian Morgan FRCPath ^9^.

**Metadata curation:**

Dr Elizabeth Wastenge MD ^53^, Dr Michael R Chapman PhD ^34,^ ^88,^ ^99^, Mr Siddharth Mookerjee MPH ^38,^ ^103^, Dr Rachael Stanley PhD ^54^, Mrs Wendy Smith ^15^, Prof Timothy Peto PhD ^59^, Dr David Eyre PhD ^59^, Dr Derrick Crook ^59^, Dr Gabrielle Vernet MBBS ^33^, Dr Christine Kitchen PhD ^10^, Huw Gulliver ^10^, Dr Ian Merrick PhD ^10^, Prof Martyn Guest PhD ^10^, Robert Munn BSc ^10^, Dr Declan T Bradley ^63,^ ^72^ and Dr Tim Wyatt ^63^.

**Project administration:**

Dr Charlotte Beaver ^99^, Luke Foulser ^99^, Sophie Palmer ^88^, Carol M Churcher ^88^, Ellena Brooks MA ^88^, Kim S Smith ^88^, Dr Katerina Galai PhD ^88^, Georgina M McManus BSc ^88^, Dr Frances Bolt PhD ^38,^ ^103^, Dr Francesc Coll PhD ^19^, Lizzie Meadows MA ^70^, Dr Stephen W Attwood PhD ^23^, Dr Alisha Davies ^69^, Elen De Lacy MSc ^69^, Fatima Downing ^69^, Sue Edwards ^69^, Dr Garry P Scarlett PhD ^76^, Mrs Sarah Jeremiah MSc ^83^ and Dr Nikki Smith PhD ^93^.

**Samples and logistics:**

Danielle Leek Bsc ^88^, Sushmita Sridhar BS ^88,^ ^99^, Sally Forrest BSc ^88^, Claire Cormie ^88^, Harmeet K Gill PhD ^88^, Joana Dias MSc ^88^, Ellen E Higginson PhD ^88^, Mailis Maes MPhil ^88^, Jamie Young BSc ^88^, Michelle Wantoch PhD ^7^, Sanger Covid Team (www.sanger.ac.uk/covid-team) ^99^, Dorota Jamrozy ^99^, Stephanie Lo ^99^, Dr Minal Patel PhD ^99^, Verity Hill ^90^, Ms Claire M Bewshea MSc ^91^, Prof Sian Ellard FRCPath ^73,^ ^91^, Dr Cressida Auckland FRCPath ^73^, Dr Ian Harrison ^66^, Dr Chloe Bishop ^66^, Dr Vicki Chalker ^66^, Dr Alex Richter PhD ^85^, Dr Andrew Beggs PhD ^85^, Dr Angus Best PhD ^86^, Dr Benita Percival PhD ^86^, Dr Jeremy Mirza PhD ^86^, Dr Oliver Megram PhD ^86^, Dr Megan Mayhew PhD ^86^, Dr Liam Crawford PhD ^86^, Dr Fiona Ashcroft PhD ^86^, Dr Emma Moles-Garcia PhD ^86^, Dr Nicola Cumley PhD ^86^, Mr Richard Hopes ^64^, Dr Patawee Asamaphan PhD ^48^, Mr Marc O Niebel MSc ^48^, Prof Rory N Gunson PhD FRCPath ^100^, Dr Amanda Bradley PhD ^52^, Dr Alasdair Maclean PhD ^52^, Dr Guy Mollett MBChB ^52^, Dr Rachel Blacow MBChB ^52^, Mr Paul Bird MSc ^16^, Mr Thomas Helmer ^16^, Miss Karlie Fallon ^16^, Dr Julian Tang ^16^, Dr Antony D Hale MBBS ^49^, Dr Louissa R Macfarlane-Smith PhD ^49^, Katherine L Harper MBiol ^49^, Miss Holli Carden MSc ^49^, Dr Nicholas W Machin MSc ^45,^ ^64^, Ms Kathryn A Jackson MSc ^92^, Dr Shazaad S Y Ahmad MSc ^45,^ ^64^, Dr Ryan P George PhD ^45^, Dr Lance Turtle PhD MRCP ^92^, Mrs Elaine O'Toole BSc ^43^, Mrs Joanne Watts BSc ^43^, Mrs Cassie Breen BSc ^43^, Mrs Angela Cowell MSc ^43^, Ms Adela Alcolea-Medina ^32,^ ^96^, Ms Themoula Charalampous MSc ^12,^ ^42^, Amita Patel ^11^, Dr Lisa J Levett PhD ^35^, Dr Judith Heaney PhD ^35^, Dr Aileen Rowan PhD ^39^, Prof Graham P Taylor DSc ^39^, Dr Divya Shah PhD ^30^, Miss Laura Atkinson MSc ^30^, Mr Jack CD Lee MSc ^30^, Mr Adam P Westhorpe BSc ^82^, Dr Riaz Jannoo PhD ^82^, Dr Helen L Lowe PhD ^82^, Miss Angeliki Karamani MSc ^82^, Miss Leah Ensell BSc ^82^, Mrs Wendy Chatterton MSc ^35^, Miss Monika Pusok MSc ^35^, Mrs Ashok Dadrah MSc ^75^, Miss Amanda Symmonds MSc ^75^, Dr Graciela Sluga MD/MSC ^44^, Dr Zoltan Molnar PhD ^72^, Mr Paul Baker MD ^79^, Prof Stephen Bonner ^79^, Ms Sarah Essex ^79^, Dr Edward Barton MD ^56^, Ms Debra Padgett BSc ^56^, Ms Garren Scott BSc ^56^, Ms Jane Greenaway MSc ^57^, Dr Brendan AI Payne MD ^50^, Dr Shirelle Burton-Fanning MD ^50^, Dr Sheila Waugh MD ^50^, Dr Veena Raviprakash MD ^17^, Ms Nicola Sheriff BSc ^17^, Ms Victoria Blakey BSc ^17^, ms Lesley-Anne Williams BSc ^17^, Dr Jonathan Moore MD ^27^, Ms Susanne Stonehouse BSc ^27^, Dr Louise Smith ^55^, Dr Rose K Davidson PhD ^89^, Dr Luke Bedford ^26^, Dr Lindsay Coupland PhD ^54^, Ms Victoria Wright BSc ^18^, Dr Joseph G Chappell PhD ^97^, Dr Theocharis Tsoleridis PhD ^97^, Prof Jonathan Ball PhD ^97^, Mrs Manjinder Khakh ^15^, Dr Vicki M Fleming PhD ^15^, Dr Michelle M Lister PhD ^15^, Dr Hannah C Howson-Wells PhD ^15^, Dr Louise Berry ^15^, Dr Tim Boswell ^15^, Dr Amelia Joseph ^15^, Dr Iona Willingham ^15^, Dr Nichola Duckworth ^60^, Dr Sarah Walsh ^60^, Dr Emma Wise PhD ^2,^ ^111^, Dr Nathan Moore PhD ^2,^ ^111^, Miss Matilde Mori BSc ^2,^ ^108,^ ^111^, Dr Nick Cortes MRCP FRCPath ^2,^ ^111^, Dr Stephen Kidd PhD ^2,^ ^111^, Dr Rebecca Williams BMBS ^33^, Laura Gifford MSc ^69^, Miss Kelly Bicknell ^61^, Dr Sarah Wyllie ^61^, Miss Allyson Lloyd ^61^, Mr Robert Impey MSc ^61^, Ms Cassandra S Malone MSc ^6^, Mr Benjamin J Cogger BSc ^6^, Nick Levene MSc ^62^, Lynn Monaghan ^62^, Dr Alexander J Keeley MRCP ^93^, Dr David G Partridge FRCP FRCPath ^78,^ ^93^, Dr Mohammad Raza ^78,^ ^93^, Dr Cariad Evans ^78,^ ^93^ and Dr Kate Johnson ^78,^ ^93^.

**Sequencing and analysis:**

Emma Betteridge BSc ^99^, Ben W Farr BSc ^99^, Scott Goodwin MSc ^99^, Dr Michael A Quail PhD ^99^, Carol Scott ^99^, Lesley Shirley MSc ^99^, Scott AJ Thurston BSc ^99^, Diana Rajan MSc ^99^, Dr Iraad F Bronner PhD ^99^, Louise Aigrain PhD ^99^, Dr Nicholas M Redshaw PhD ^99^, Dr Stefanie V Lensing PhD ^99^, Shane McCarthy ^99^, Alex Makunin ^99^, Dr Carlos E Balcazar PhD ^90^, Dr Michael D Gallagher PhD ^90^, Dr Kathleen A Williamson PhD ^90^, Thomas D Stanton BSc ^90^, Ms Michelle L Michelsen BSc ^91^, Ms Joanna Warwick-Dugdale BSc ^91^, Dr Robin Manley Ph.D. ^91^, Ms Audrey Farbos MSc ^91^, Dr James W Harrison Ph.D. ^91^, Dr Christine M Sambles Ph.D. ^91^, Dr David J Studholme PhD. ^91^, Dr Angie Lackenby ^66^, Dr Tamyo Mbisa ^66^, Dr Steven Platt ^66^, Mr Shahjahan Miah ^66^, Dr David Bibby ^66^, Dr Carmen Manso ^66^, Dr Jonathan Hubb ^66^, Dr Gavin Dabrera ^66^, Dr Mary Ramsay ^66^, Dr Daniel Bradshaw ^66^, Dr Ulf Schaefer ^66^, Dr Natalie Groves ^66^, Dr Eileen Gallagher ^66^, Dr David Lee ^66^, Dr David Williams ^66^, Dr Nicholas Ellaby ^66^, Hassan Hartman ^66^, Nikos Manesis ^66^, Vineet Patel ^66^, Juan Ledesma ^67^, Ms Katherine A Twohig ^67^, Dr Elias Allara ^64,^ ^88^, Ms Clare Pearson ^64,^ ^88^, Mr Jeffrey K. J. Cheng MSc ^94^, Dr Hannah E. Bridgewater PhD ^94^, Ms Lucy R. Frost BSc ^94^, Ms Grace Taylor-Joyce BSc ^94^, Dr Paul E Brown PhD ^94^, Dr Lily Tong PhD ^48^, Ms Alice Broos BSc ^48^, Mr Daniel Mair BSc ^48^, Mrs Jenna Nichols BSc ^48^, Dr Stephen N Carmichael PhD ^48^, Dr Katherine L Smollett PhD ^40^, Dr Kyriaki Nomikou PhD ^48^, Dr Elihu Aranday-Cortes PhD/DVM ^48^, Ms Natasha Johnson BSc ^48^, Dr Seema Nickbakhsh PhD ^48,^ ^68^, Dr Edith E Vamos PhD ^92^, Dr Margaret Hughes PhD ^92^, Dr Lucille Rainbow PhD ^92^, Mr Richard Eccles MSc ^92^, Ms Charlotte Nelson MSc ^92^, Dr Mark Whitehead PhD ^92^, Dr Richard Gregory PhD ^92^, Mr Matthew Gemmell MSc ^92^, Ms Claudia Wierzbicki BSc ^92^, Ms Hermione J Webster BSc ^92^, Ms Chloe L Fisher MSc ^28^, Mr Adrian W Signell BSc ^20^, Dr Gilberto Betancor PhD ^20^, Mr Harry D Wilson BSc ^20^, Dr Gaia Nebbia PhD FRCPath ^12^, Dr Flavia Flaviani PhD ^31^, Mr Alberto C Cerda MSc ^96^, Ms Tammy V Merrill MSc ^96^, Rebekah E Wilson MSc ^96^, Mr Marius Cotic MSc ^82^, Miss Nadua Bayzid BSc ^82^, Dr Thomas Thompson PhD ^72^, Dr Erwan Acheson PhD ^72^, Prof Steven Rushton PhD ^51^, Prof Sarah O'Brien PhD ^51^, David J Baker BEng ^70^, Steven Rudder ^70^, Alp Aydin MSci ^70^, Dr Fei Sang PhD ^18^, Dr Johnny Debebe PhD ^18^, Dr Sarah Francois PhD ^23^, Dr Tetyana I Vasylyeva DPhil ^23^, Dr Marina Escalera Zamudio PhD ^23^, Mr Bernardo Gutierrez MSc ^23^, Dr Angela Marchbank BSc ^10^, Joshua Maksimovic FD ^9^, Karla Spellman FD ^9^, Kathryn McCluggage Msc ^9^, Dr Mari Morgan PhD ^69^, Robert Beer BSc ^9^, Safiah Afifi BSc ^9^, Trudy Workman HNC ^10^, William Fuller BSc ^10^, Catherine Bresner Bsc ^10^, Dr Adrienn Angyal PhD ^93^, Dr Luke R Green PhD ^93^, Dr Paul J Parsons PhD ^93^, Miss Rachel M Tucker MSc ^93^, Dr Rebecca Brown PhD ^93^ and Mr Max Whiteley PhD ^93^

**Software and analysis tools:**

James Bonfield BSc ^99^, Dr Christoph Puethe ^99^, Mr Andrew Whitwham BSc ^99^, Jennifier Liddle ^99^, Dr Will Rowe PhD ^41^, Dr Igor Siveroni PhD ^39^, Dr Thanh Le-Viet PhD ^70^ and Amy Gaskin MSc ^69^.

**Visualisation:**

Dr Rob Johnson PhD ^39^.

**1** Barking, Havering and Redbridge University Hospitals NHS Trust, **2** Basingstoke Hospital, **3** Belfast Health & Social Care Trust, **4** Betsi Cadwaladr University Health Board, **5** Big Data Institute, Nuffield Department of Medicine, University of Oxford, **6** Brighton and Sussex University Hospitals NHS Trust, **7** Cambridge Stem Cell Institute, University of Cambridge, **8** Cambridge University Hospitals NHS Foundation Trust, **9** Cardiff and Vale University Health Board, **10** Cardiff University, **11** Centre for Clinical Infection & Diagnostics Research, St. Thomas' Hospital and Kings College London, **12** Centre for Clinical Infection and Diagnostics Research, Department of Infectious Diseases, Guy's and St Thomas' NHS Foundation Trust, **13** Centre for Enzyme Innovation, University of Portsmouth (PORT), **14** Centre for Genomic Pathogen Surveillance, University of Oxford, **15** Clinical Microbiology Department, Queens Medical Centre, **16** Clinical Microbiology, University Hospitals of Leicester NHS Trust, **17** County Durham and Darlington NHS Foundation Trust, **18** Deep Seq, School of Life Sciences, Queens Medical Centre, University of Nottingham, **19** Department of Infection Biology, Faculty of Infectious & Tropical Diseases, London School of Hygiene & Tropical Medicine, **20** Department of Infectious Diseases, King's College London, **21** Department of Microbiology, Kettering General Hospital, **22** Departments of Infectious Diseases and Microbiology, Cambridge University Hospitals NHS Foundation Trust; Cambridge, UK, **23** Department of Zoology, University of Oxford, **24** Division of Virology, Department of Pathology, University of Cambridge, **25** East Kent Hospitals University NHS Foundation Trust, **26** East Suffolk and North Essex NHS Foundation Trust, **27** Gateshead Health NHS Foundation Trust, **28** Genomics Innovation Unit, Guy's and St. Thomas' NHS Foundation Trust, **29** Gloucestershire Hospitals NHS Foundation Trust, **30** Great Ormond Street Hospital for Children NHS Foundation Trust, **31** Guy's and St. Thomas’ BRC, **32** Guy's and St. Thomas’ Hospitals, **33** Hampshire Hospitals NHS Foundation Trust, **34** Health Data Research UK Cambridge, **35** Health Services Laboratories, **36** Heartlands Hospital, Birmingham, **37** Hub for Biotechnology in the Built Environment, Northumbria University, **38** Imperial College Hospitals NHS Trust, **39** Imperial College London, **40** Institute of Biodiversity, Animal Health & Comparative Medicine, **41** Institute of Microbiology and Infection, University of Birmingham, **42** King's College London, **43** Liverpool Clinical Laboratories, **44** Maidstone and Tunbridge Wells NHS Trust, **45** Manchester University NHS Foundation Trust, **46** Microbiology Department, Wye Valley NHS Trust, Hereford, **47** MRC Biostatistics Unit, University of Cambridge, **48** MRC-University of Glasgow Centre for Virus Research, **49** National Infection Service, PHE and Leeds Teaching Hospitals Trust, **50** Newcastle Hospitals NHS Foundation Trust, **51** Newcastle University, **52** NHS Greater Glasgow and Clyde, **53** NHS Lothian, **54** Norfolk and Norwich University Hospital, **55** Norfolk County Council, **56** North Cumbria Integrated Care NHS Foundation Trust, **57** North Tees and Hartlepool NHS Foundation Trust, **58** Northumbria University, **59** Oxford University Hospitals NHS Foundation Trust, **60** PathLinks, Northern Lincolnshire & Goole NHS Foundation Trust, **61** Portsmouth Hospitals University NHS Trust, **62** Princess Alexandra Hospital Microbiology Dept., **63** Public Health Agency, **64** Public Health England, **65** Public Health England, Clinical Microbiology and Public Health Laboratory, Cambridge, UK, **66** Public Health England, Colindale, **67** Public Health England, Colindale, **68** Public Health Scotland, **69** Public Health Wales NHS Trust, **70** Quadram Institute Bioscience, **71** Queen Elizabeth Hospital, **72** Queen's University Belfast, **73** Royal Devon and Exeter NHS Foundation Trust, **74** Royal Free NHS Trust, **75** Sandwell and West Birmingham NHS Trust, **76** School of Biological Sciences, University of Portsmouth (PORT), **77** School of Pharmacy and Biomedical Sciences, University of Portsmouth (PORT), **78** Sheffield Teaching Hospitals, **79** South Tees Hospitals NHS Foundation Trust, **80** Swansea University, **81** University Hospitals Southampton NHS Foundation Trust, **82** University College London, **83** University Hospital Southampton NHS Foundation Trust, **84** University Hospitals Coventry and Warwickshire, **85** University of Birmingham, **86** University of Birmingham Turnkey Laboratory, **87** University of Brighton, **88** University of Cambridge, **89** University of East Anglia, **90** University of Edinburgh, **91** University of Exeter, **92** University of Liverpool, **93** University of Sheffield, **94** University of Warwick, **95** University of Cambridge, **96** Viapath, Guy's and St Thomas' NHS Foundation Trust, and King's College Hospital NHS Foundation Trust, **97** Virology, School of Life Sciences, Queens Medical Centre, University of Nottingham, **98** Wellcome Centre for Human Genetics, Nuffield Department of Medicine, University of Oxford, **99** Wellcome Sanger Institute, **100** West of Scotland Specialist Virology Centre, NHS Greater Glasgow and Clyde, **101** Department of Medicine, University of Cambridge, **102** Ministry of Health, Sri Lanka, **103** NIHR Health Protection Research Unit in HCAI and AMR, Imperial College London, **104** North West London Pathology, **105** NU-OMICS, Northumbria University, **106** University of Kent, **107** University of Oxford, **108** University of Southampton, **109** University of Southampton School of Health Sciences, **110** University of Southampton School of Medicine, **111** University of Surrey, **112** Warwick Medical School and Institute of Precision Diagnostics, Pathology, UHCW NHS Trust, **113** Wellcome Africa Health Research Institute Durban and **114** Wellcome Genome Campus.
